# Observational study of effects of HIV Acquisition and Antiretroviral Treatment on Biomarkers of Systemic Immune Activation

**DOI:** 10.1101/2023.07.07.23292352

**Authors:** Ewelina Kosmider, Jackson Wallner, Ana Gervassi, Rachel A Bender Ignacio, Delia Pinto-Santini, German Gornalusse, Urvashi Pandey, Florian Hladik, Paul T. Edlefsen, Javier R. Lama, Ann C. Duerr, Lisa M. Frenkel

## Abstract

**Objective:** Assess whether biomarkers of systemic inflammation are associated with HIV acquisition or with the timing of ART initiation (“immediate”, at diagnosis, versus “deferred”, at 24 weeks postdiagnosis) in men-who-have-sex-with-men (MSM) and transgender women.

**Design:** A retrospective study comparing inflammatory biomarkers in participants’ specimens collected before and after ≥2 years of effective ART.

**Methods:** Inflammatory biomarkers were measured in four longitudinally collected plasma specimens, including two plasma specimens collected from each participant before and two after HIV acquisition and confirmed ART-suppression. Biomarkers were quantified by enzyme-linked immuno-assay or Meso Scale Discovery. Statistical measures compared intra-participant and between-group changes in biomarkers.

**Results:** Across 50 participants, the levels of C-reactive protein (CRP), monocyte chemo-attractant protein-1, tumor necrosis factor-α and interferon gamma-induced protein-10 significantly increased while leptin and lipopolysaccharide binding protein (LBP) significantly decreased following HIV infection. Randomization to deferred-ART initiation was associated with greater increases in CRP and no decreases in LBP. Multiple biomarkers varied significantly within participants’ two pre-infection or two post-ART-suppression specimens.

**Conclusions:** Acquisition of HIV appeared to induce systemic inflammation, with elevation of biomarkers previously associated with infections and cardiovascular disease. Initiation of ART during the early weeks of infection tempered the increase in pro-inflammatory biomarkers compared to those who delayed ART for ∼24 weeks after HIV diagnosis, perhaps because immediate-ART limited the size of the HIV reservoir or limited immune dysregulation. Some but not all biomarkers appeared sufficiently stable to assess intraparticipant changes over time. Given that pro-inflammatory biomarkers predict multiple co-morbidities, our findings suggest that immediate-ART initiation may improve health outcomes.

## Introduction

The progression of HIV disease to AIDS has been mitigated by antiretroviral treatment (**ART**) (1, 2, 3). Despite ART, life-threatening non-AIDS-defining comorbidities associated with elevated biomarkers of systemic immune activation occur at increased frequencies in people living with HIV (**PWH**) compared to uninfected persons (4, 5, 6, 7, 8, 9, 10, 11, 12, 13, 14). While effective ART is associated with normalization of some pro-inflammatory biomarkers, other biomarkers, particularly those associated with monocyte/macrophage activation, remain elevated in PWH compared to uninfected individuals (7). Studies comparing immune activation biomarkers in PWH versus uninfected persons aim to control for genetic, behavioral, or other pre-existing factors (4, 7, 15). However, longitudinal studies examining these factors before and following incident HIV infection are lacking. We leveraged specimens collected from individuals with prospectively documented incident HIV infection (16, 17) to compare immune biomarkers in specimens collected prior to HIV infection to those collected after ART suppression of viral replication. Because ART prevents ongoing viral replication but does not prevent infected cells from producing viral proteins and particles, we hypothesized that PWH would demonstrate greater immune activation after ART suppression of viral replication compared to pre-infection values. Additionally, because ART initiated during primary infection limits the size of the HIV reservoir, we hypothesized that PWH initiating ART at diagnosis during acute or early primary infection (“immediate” ART) would show less immune activation compared to those who “deferred” ART for ∼24 weeks.

## METHODS

Banked specimens from a prospective study of incident HIV infection among seronegative men-who-have-sex-with-men (**MSM**) and transgender women in Lima, Peru (*Sabes* Study) (16) were utilized to evaluate immune biomarkers. Participants were enrolled into the *Sabes* Study between 7/16/2013 and 7/31/2015 followed through 9/10/2019 (date of last 4^th^ timepoint sample). All participants provided written informed consent including consent for future use of specimens; personal identifiers were retained at the study site in Lima (JRL) for participant tracking and were not available to other co-authors.

Participants were screened monthly for HIV acquisition by HIV antibody and nucleic acid amplification testing. The estimated date of detectable infection (**EDDI**) was calculated as previously described (17, 18). Following HIV diagnosis, participants were randomized to initiate ART immediately (**immediate-ART**) or to defer ART for 24 weeks (**deferred-ART**) (16).

Participants with documented incident HIV infection were selected for this sub-study in February 2020 (with participants data accessed on multiple dates during the preceding months) based on availability of two pre-and two post-infection plasma specimens, with the rationale to evaluate immune biomarkers in steady-state and avoid transient changes associated with HIV acquisition or ART initiation. Specimens included one plasma sample shortly after enrollment, a second ≤3 months from EDDI (**Visits 1** and **2**, respectively), a third ≥6 months and a fourth ≥24 months after ART suppression of plasma HIV RNA to <200 copies/mL (ART-suppression) (**Visits 3** and **4**, respectively).

Biomarkers linked to systemic inflammation and cardiovascular disease were selected for quantification: C-reactive protein (**CRP**), tumor necrosis factor-α (**TNF-α**), interleukin (**IL-6**), soluble urokinase-type plasminogen activator receptor (**suPAR**), interferon gamma-induced protein 10 (**IP-10**), interleukin 1β (**IL-1β)**, interleukin 8 **(IL-8)**^7^, interleukin 10 **(IL-10**) (19, 20, 21), lipopolysaccharide binding protein (**LBP**)) (22, 23), and markers associated with bacterial translocation (soluble cluster of differentiation 14 and 163 (**sCD14, sCD163**), and antiviral responses (interferon-gamma (**IFN-γ**), leptin, interleukin α-2a (**IFN-α2a)**, monocyte chemoattractant protein-1 (**MCP-1/CCL2**)) (24). Meso Scale Discovery (**MSD**, Rockville, MD) determined levels of IFN-α2a, IFN-γ, IL-1β, IL-6, IL-8, IL-10, IP-10, leptin, MCP-1/CCL2, TNF-α, LBP, CRP, and ELISAs (R&D Systems, Minneapolis, MN) determined levels of sCD14, sCD163, and suPAR in March/April 2020.

During study analysis, participants’ data was accessed on multiple dates in November and December 2020 and throughout 2021 by multiple team members. To determine the stability of biomarkers across the two pre-infection or two post-suppression time-points, a two-sided, one-sample t-test was used to separately compare values within each pair of pre-infection and post-suppression specimens. Biomarker levels were also compared to physiologic “normal” ranges determined by assay manufacturers or clinical studies (leptin, CRP, IFN-α2a) (25, 26). Because a prior study associated elevated plasma sCD14 levels with efavirenz-based-ART(8), we further analyzed values across Visit-3 and -4 in participants switching from efavirenz-to non-efavirenz-based regimens.

To identify biomarkers that differed following HIV infection despite more than six months of ART-suppression, a regression analysis was conducted separately for each biomarker. Participant-specific fixed effects were evaluated across timeframes defined by pre-or post-infection paired visits. To account for variations within the pre- and within the post-infection specimens, variance was pooled across timeframes and across people. This effectively allowed us to implement a pooled variance one-sample t-test with repeated measures, treating the paired visits within the pre-or post-infection timeframe as exchangeable. Unadjusted and Holm adjusted p-values and 95% confidence intervals for the change in mean marker value pre-versus post-infection were calculated in R 4.2.1. Additionally, the regression model included the indicator “deferred-ART” to evaluate whether the duration of infection prior to ART initiation contributed to a difference in biomarkers. P-values of ≤0.05 were considered significant. To display multiple biomarker differences on a common scale, the differences between pre-versus post-infection means were divided by the pre-infection standard deviation (**SD**) for each biomarker.

## RESULTS

### Participants

From among a total of 216 *Sabes* participants with prospectively documented incident HIV infection 50 had specimens from four time-points and fulfilled ART-suppression entry criteria and were used for this study of immune biomarkers. These 50 included 19 participants randomized to immediate-ART and 31 randomized to deferred-ART. Five participants randomized to deferred-ART initiated ART prior to 24-weeks post-HIV-diagnosis due to low CD4 cell counts or other ART-qualifying events and two of these initiated ART in the immediate-ART timeframe and are included in the immediate-ART group for the “as-treated” analysis; three initiated ART between the immediate- and the deferred-ART timeframes and The antiretrovirals provided to study participants shifted over time: at Visit 3, 43/50 were receiving efavirenz+emtricitabine+tenofovir disoproxil and by Visit 4, 48/50 were receiving elvitegravir/cobicistat+emtricitabine+tenofovir alafenamide. Non-study ART regimens were given to nine participants as medically indicated, including seven protease-inhibitor-based and two efavirenz-based regimens.

### Pro-inflammatory biomarkers

Prior to HIV infection the paired pre-infection biomarker levels demonstrated intraparticipant stability, except for significant variability in IP-10, IL-6 and sCD163 (N=50, **Supplementary Table 1, Supplementary Figure 1**). The mean biomarker values for most participants were within established normal ranges (**Figure 1**). Outliers with elevated biomarkers observed in both pre-infection specimens included IL-1β (N=1 participant), suPAR (N=2), MCP-1/CCL2, sCD163, and CRP (N=3), TNFα (N=9) and/or leptin (N=23). Following ART-suppression, intraparticipant biomarkers demonstrated stability except for sCD163, leptin, IL-8, and LBP (N=50, **Supplementary Table 2, Supplementary Figure 2**). The mean biomarker levels of most participants remained within established norms, except outliers with both values elevated were observed for IL-1β (N=1 participant), suPAR and sCD163 (N=4), IL-6 (N=5), leptin (N=6), MCP-1/CCL2 (N=8), CRP and TNF-α (N=9) (**Figure 1**).

**Table 1:**
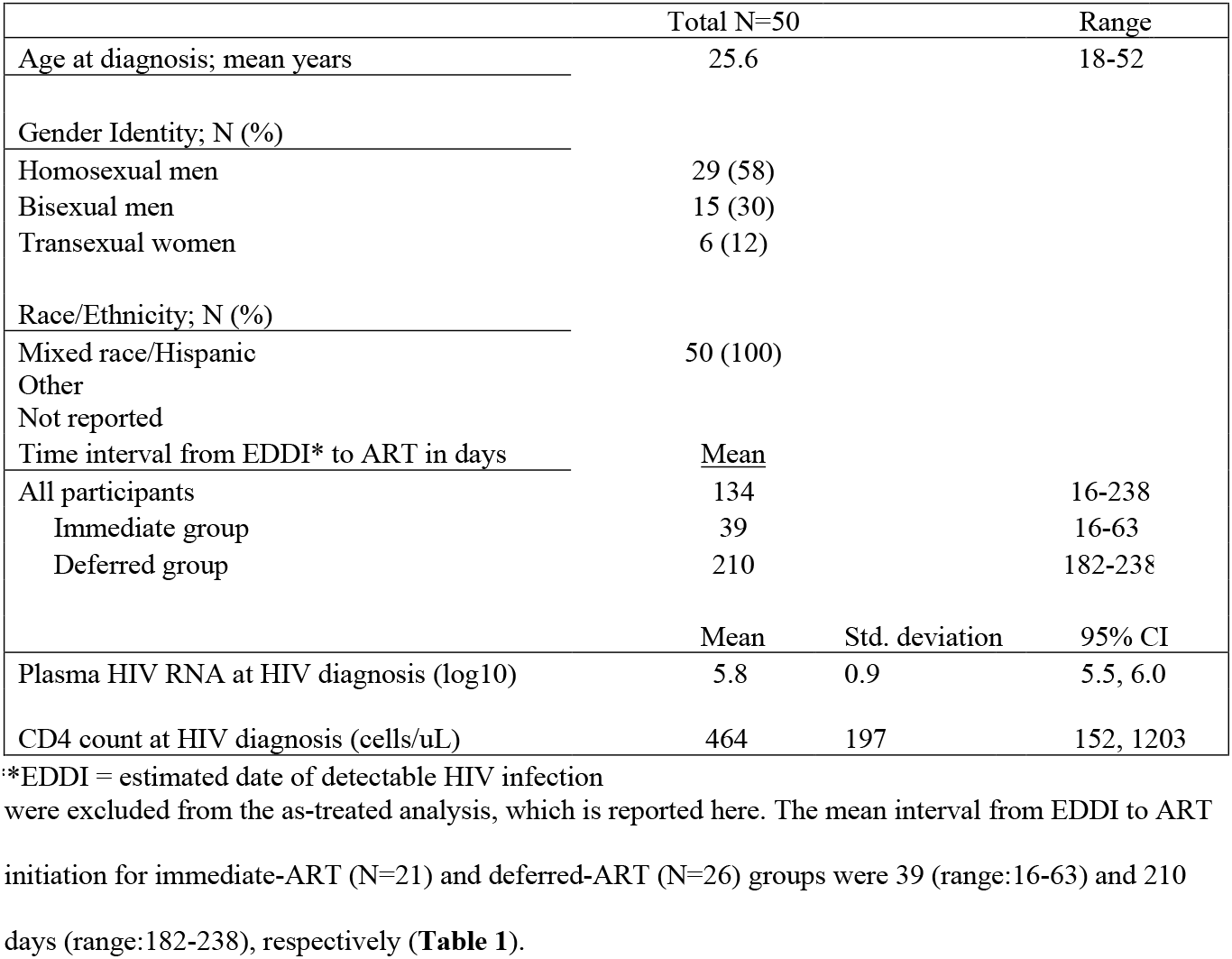
Demographic and clinical parameters of participants

|  | Total N=50 |  | Range |
| --- | --- | --- | --- |
| Age at diagnosis; mean years | 25.6 |  | 18-52 |
| Gender Identity; N (%) |  |  |  |
| Homosexual men | 29 (58) |  |  |
| Bisexual men | 15 (30) |  |  |
| Transsexual women | 6 (12) |  |  |
| Race/Ethnicity; N (%) |  |  |  |
| Mixed race/Hispanic | 50 (100) |  |  |
| Other |  |  |  |
| Not reported |  |  |  |
| Time interval from EDDI* to ART in days | <u>Mean</u> |  |  |
| All participants | 134 |  | 16-238 |
| Immediate group | 39 |  | 16-63 |
| Deferred group | 210 |  | 182-238 |
|  | Mean | Std. deviation | 95% CI |
| Plasma HIV RNA at HIV diagnosis (log10) | 5.8 | 0.9 | 5.5, 6.0 |
| CD4 count at HIV diagnosis (cells/uL) | 464 | 197 | 152, 1203 |

\*EDDI = estimated date of detectable HIV infection

**Figure 1.**
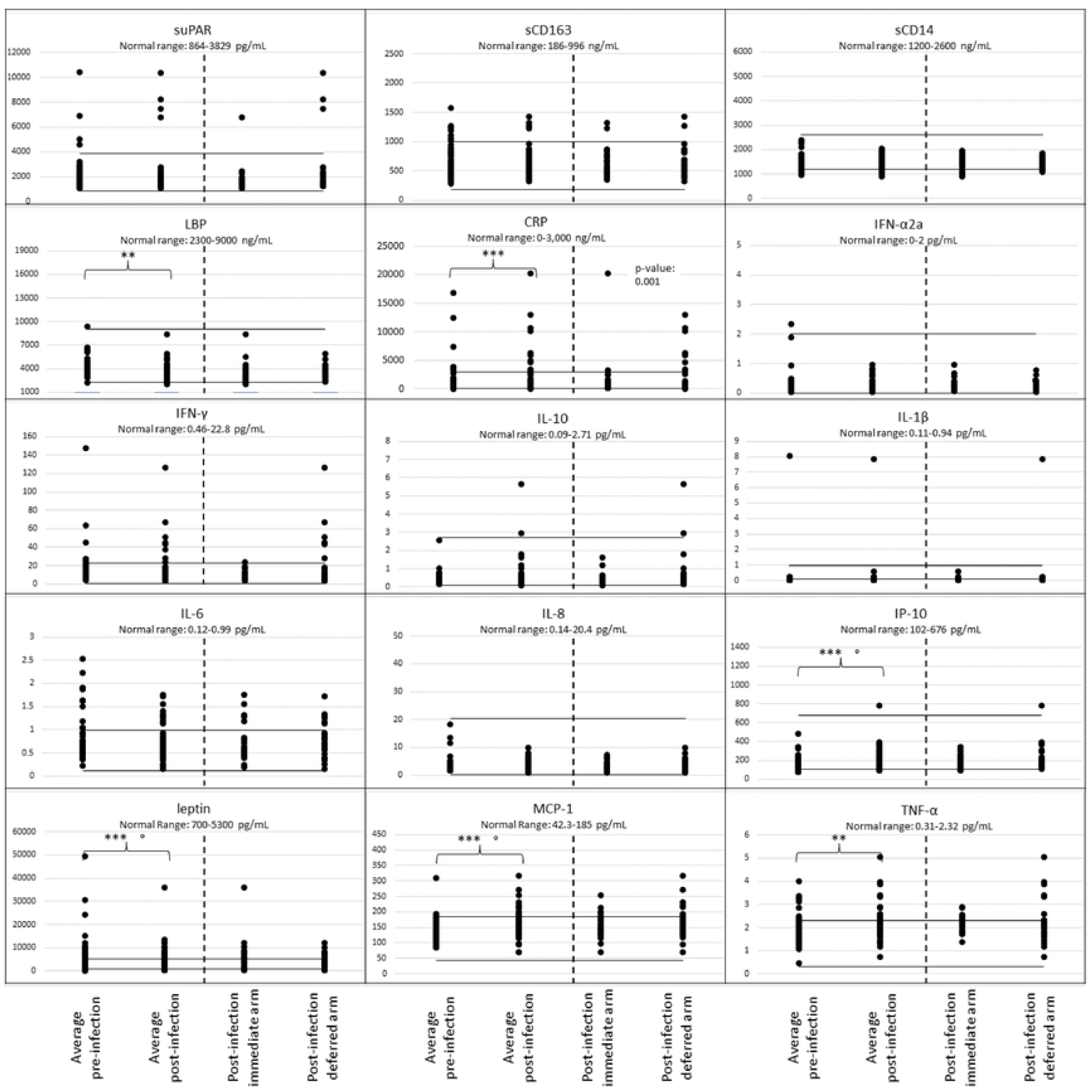
Plasma biomarker levels comparing pre-HIV-infection to post-infection and antiretroviral therapy-suppression by timing of ART initiation. Each biomarker evaluated is shown in a separate panel, with normal upper and lower ranges indicated by solid horizontal lines. The mean biomarker levels are plotted for each participant’s (N=50) biomarker values from two timepoints prior to HIV infection (pre-acquisition) and two timepoints post-ART-suppression (>6 months and >2 years post ART suppression of plasma HIV RNA to <200 c/mL). The two post-ART-suppression values also shown for “all” participants and separately by the two “as-treated” groups by the time when ART was initiated, either immediately upon diagnosis during primary infection (N=21) or deferred for 24 weeks after HIV diagnosis (N=26) (Immediate-ART, Deferred-ART, respectively). Unadjusted p-values by a regression analysis shown (* <0.01, ** < 0.05, *** < 0.001) for significant changes in mean cytokine levels between the 47 participants prior to HIV infection versus after ART-suppression. Holm adjusted p-values <0.001 indicated by (°).

Comparisons of all participant’s (N=47) mean pre-infection biomarker values to their ART-suppressed mean values by a regression analysis detected statistically significant increases in IP-10, MCP-1/CCL2, TNFα, CRP and significant decreases in leptin and LBP (**Figure 1**), with differences sustained after Holm adjustment for multiple comparisons in all but LBP and TNF-α (**Supplementary Table 3a**). Comparison of biomarker levels by timing of ART-initiation found a difference in pre-infection to post-ART-suppression by ART timing group for IFN-α2a and CRP (**Figure 2** and **Supplementary Tables 4a**); CRP increased and IFN-α2a decreased in the deferred-ART but not in the immediate-ART group (**Figure 2, Supplementary Table 5a, Supplementary Table 6a**). Furthermore, IP-10 and MCP-1/CCL2 increased and leptin decreased in both groups, while LBP decreased in the immediate-ART group but not in the deferred-ART group.

**Figure 2.**
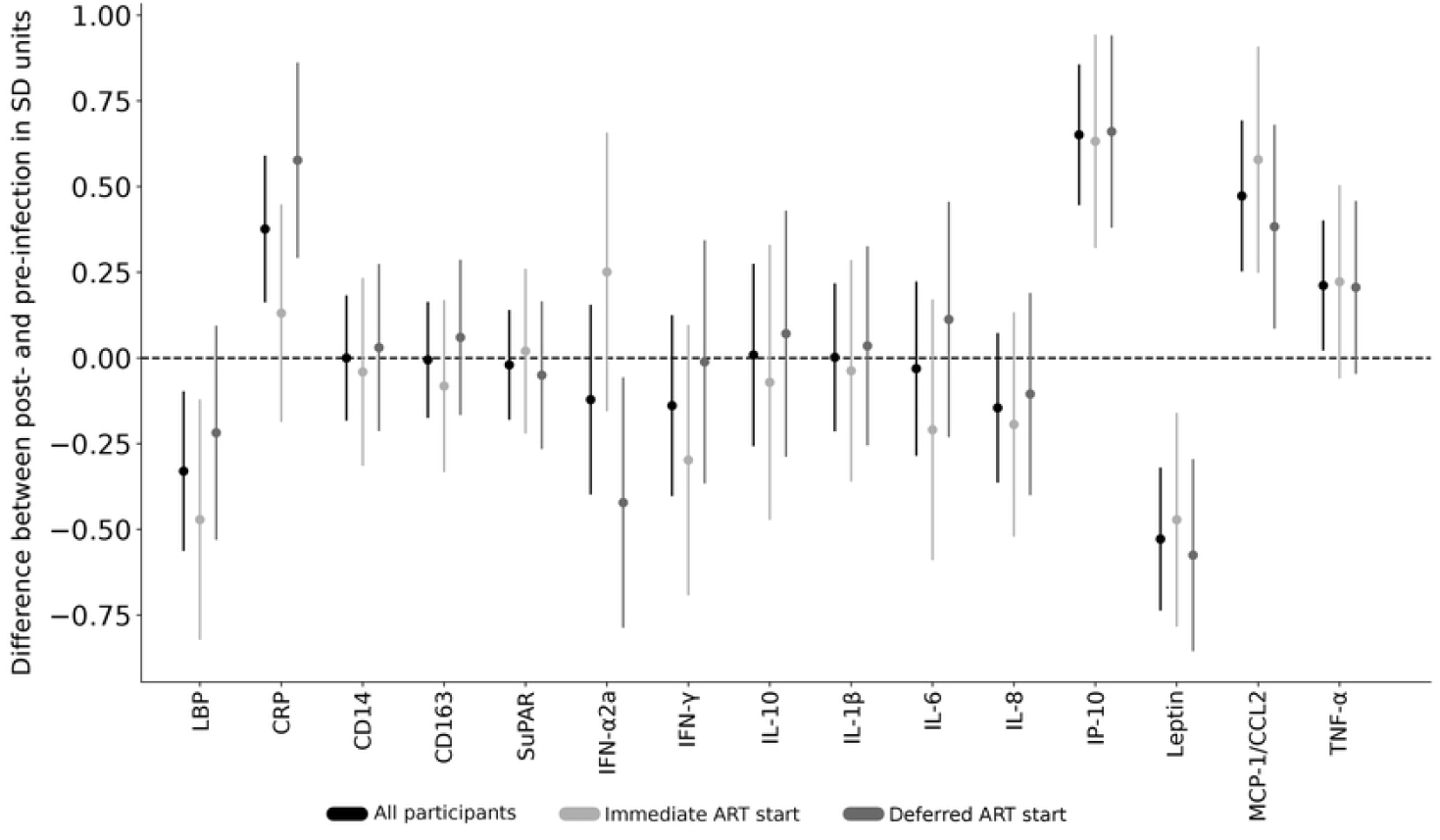
Difference between pre-HIV-infection and post-ART suppression biomarker values by timing of antiretroviral therapy (ART) initiation. Biomarker levels from two specimens before HIV infection and two after ART-suppression were separately compared for all participants (shown in black (N=47)), and participants separated into two groups: those who started ART immediately upon HIV-diagnosis in light gray (N=21) vs. those who deferred ART initiation for 24 weeks in dark gray (N=26), with mean differences (dots) and 95% confidence intervals (lines) shown. The 95% confidence intervals were calculated based on a regression analysis with values log transformed prior to analysis and differences divided by pre-infection standard deviation for each analyte. The vertical dotted line reflects no difference between post- and pre-infection values. Abbreviations: suPAR, soluble urokinase-type plasminogen activator receptor; sCD14 and sCD163, soluble cluster of differentiation 14 and 163; LBP, lipopolysaccharide binding protein; IL-1β, IL-6, IL-8 and IL-10, interleukin 1b, 6, 8 and 10; IFN-γ and IFN-α2a, interferon-gamma and -alpha 2a; IP-10, interferon gamma-induced protein 10; MCP-1/CCL2, monocyte chemoattractant protein-1; TNF-α, tumor necrosis factor-alpha; CRP, C-reactive protein A comparison of sCD14 values between Visit-3 and -4 in participants (N=43) who switched from efavirenz-to non-efavirenz-based regimens found no significant changes between these intervals.

## DISCUSSION

This study is unique in documenting changes in biomarkers of immune activation in individuals with prospectively documented incident HIV infection, and in examining differences in biomarkers between participants randomized to initiate ART immediately in early infection or to defer ART for 24 weeks. Comparison of biomarkers from pre-infection to post ART-suppression, including correction for multiple comparisons, showed increased plasma levels of proinflammatory chemokines/cytokines MCP-1/CCL2 and IP-10 secreted by monocyte/macrophages and other cell types in response to HIV infection (27, 28) or stimulation by cytokines (29), and CRP, a marker of inflammation associated with infections, cancers, auto-immunity or tissue damage. These findings of elevated inflammatory cytokines/chemokines are consistent with those reported by previous studies (4, 6, 7, 30). In addition, the levels of one biomarker, leptin, were lower in the post-infection compared to pre-infection specimens. The decrease in leptin, a marker of energy expenditure (31), is consistent with previous studies finding lower leptin in ART-treated PWH (32), likely due to HIV-infection-induced catabolism.

Multiple biomarkers, including IP-10, IL-6, sCD163, leptin, IL-8, and LBP were variable either within each participant’s two pre-infection or two post-ART-suppression specimens. Temporal perturbations in these analytes may have occurred due to intercurrent illnesses, alcohol consumption (33, 34), or other unknown reasons. The observed intraparticipant variation suggests that a determination of these analytes at a single point in time may be unreliable in assessing an individual’s biomarker levels and supports the use of large datasets to explore relationships between HIV infection and markers of immune activation.

Our comparison of biomarker levels between those who initiated immediate- versus deferred-ART initiation found that those treated immediately had significantly less elevation of their CRP and LBP values. Earlier initiation of ART limits the size of the persistent viral reservoir (35), which should limit production of viral nucleic acids and proteins that others have found associated with progression of carotid artery intima thickness (13) or atherosclerotic plaque (14). The greater decrease in plasma IFN-α2a observed among those deferring ART (**Figure 2**) may be attributable to consistently high pre-infection values in this group. Notably, during ART all participants had IFN-α2a values in the normal range of the assay (**Figure 1**).

While efavirenz in prior studies was associated with elevated sCD14 and kynurenine-tryptophan ratio (8, 36), we did not observe a significant change in sCD14 in participants who switched from efavirenz-based ART to a “non-efavirenz” elvitegravir-based regimen. It is not known whether elvitegravir (or cobicistat, contained in the co-formulated product to reduce hepatic clearance of elvitegravir) is associated with inflammation, but it is notable that sCD14 levels were in the normal range for all specimens tested in the study.

The primary limitations of this study are the relatively small size of the cohort examined, a relatively short follow-up of the ART-suppressed participants for this life-long infection and the assessment of biomarkers from relatively few timepoints. In addition, the relative youth of our study participants and the short duration of their HIV infections limited our ability to observe non-AIDS adverse events, and we were unable to conduct long term follow-up to observe and correlate our findings with clinical events. Additionally, when evaluating the potential impact of efavirenz, we did not test some biomarkers found to be abnormal in other studies, e.g., kynurenine/tryptophan ratio (8, 36). The primary strength of this study comes from the comparison of two samples from before and two after documented incident HIV infection. The two specimens prior and two after infection reduces variability due to extraneous events.

The study of individuals with incident infection diminishes the biases due to pre-existing conditions and confounding behavioral practices, although, we acknowledge that behaviors may change following HIV diagnosis (37).

## CONCLUSIONS

In conclusion, multiple pro-inflammatory biomarkers appear to have been induced by HIV and/or ART, despite virologic suppression. Importantly, ART initiation during acute/early HIV infection appeared to limit CRP levels. Given the strong association of CRP with cardiovascular disease (38, 39), these findings emphasize that HIV prevention and ART initiation during primary infection could diminish non-AIDS events.

## Data Availability

Summary data available in the manuscript. Participant level data is stored at Fred Hutch and de-identified data is available by request of Ann Duerr

## ACKNOWLEDGEMENTS and FUNDING

The authors acknowledge the contributions of the study participants, site investigators and staff. The study drug was provided at no cost to the Institution by Merck Sharp & Dohme Corp. The opinions expressed in this paper are those of the authors and do not necessarily represent those of Merck Sharp & Dohme Corp.

The project was funded by NIH R01 DA040532 (ACD). Additional support came from the Molecular Profiling and Computational Biology Core of the University of Washington and Fred Hutch Center for AIDS Research (award number P30 AI027757), from the Laboratory Core of International Maternal Pediatric Adolescent AIDS Clinical Trials Network (IMPAACT) UM1 AI106716 (Subaward: LMF). This work was also supported by the National Center For Advancing Translational Sciences of the National Institutes of Health under Award Number KL2 TR002317 (GG). The content is solely the responsibility of the authors and does not necessarily represent the official views of the National Institutes of Health.

## SEQUENCE DATA

Nucleotide consensus sequences are available in the NCBI Genbank under accession numbers (pending manuscript acceptance). Illumina data are available in the NCBI Sequence Read Archive under BioProject number (pending manuscript acceptance).

## REFERENCES

1. Fischl MA, Richman DD, Grieco MH, Gottlieb MS, Volberding PA, Laskin OL, et al. The efficacy of azidothymidine (AZT) in the treatment of patients with AIDS and AIDS-related complex. New England Journal of Medicine. 1987;317(4):185–91.

2. Paterson DL, Swindells S, Mohr J, Brester M, Vergis EN, Squier C, et al. Adherence to protease inhibitor therapy and outcomes in patients with HIV infection. Annals of internal medicine. 2000;133(1):21–30.

3. Zolopa AR, Andersen J, Komarow L, Sanne I, Sanchez A, Hogg E, et al. Early antiretroviral therapy reduces AIDS progression/death in individuals with acute opportunistic infections: a multicenter randomized strategy trial. PloS one. 2009;4(5):e5575.

4. Hellmuth J, Slike BM, Sacdalan C, Best J, Kroon E, Phanuphak N, et al. Very early initiation of antiretroviral therapy during acute HIV infection is associated with normalized levels of immune activation markers in cerebrospinal fluid but not in plasma. The Journal of infectious diseases. 2019;220(12):1885–91.

5. Sunil M, Nigalye M, Somasunderam A, Martinez ML, Yu X, Arduino RC, et al. Unchanged levels of soluble CD14 and IL-6 over time predict serious non-AIDS events in HIV-1-infected people. AIDS research and human retroviruses. 2016;32(12):1205–9.

6. Sereti I, Krebs SJ, Phanuphak N, Fletcher JL, Slike B, Pinyakorn S, et al. Editor’s choice: Persistent, Albeit Reduced, Chronic Inflammation in Persons Starting Antiretroviral Therapy in Acute HIV Infection. Clinical infectious diseases: an official publication of the Infectious Diseases Society of America. 2017;64(2):124.

7. Wada NI, Jacobson LP, Margolick JB, Breen EC, Macatangay B, Penugonda S, et al. The effect of HAART-induced HIV suppression on circulating markers of inflammation and immune activation. AIDS (London, England). 2015;29(4):463.

8. Schnittman SR, Deitchman AN, Beck-Engeser G, Ahn H, York VA, Hartig H, et al. Abnormal Levels of Some Biomarkers of Immune Activation Despite Very Early Treatment of Human Immunodeficiency Virus. The Journal of Infectious Diseases. 2021;223(9):1621–30.

9. Lundgren JD, Babiker AG, Gordin F, Emery S, Grund B, Sharma S, et al. Initiation of antiretroviral therapy in early asymptomatic HIV infection. The New England journal of medicine. 2015;373(9):795–807.

10. Temprano ANS 12136 Study Group, Danel C, Moh R, et al. A trial of early antiretrovirals and isoniazid preventive therapy in Africa. New England Journal of Medicine. 2015;373(9):808–22.

11. Rasmussen LD, May MT, Kronborg G, Larsen CS, Pedersen C, Gerstoft J, et al. Time trends for risk of severe age-related diseases in individuals with and without HIV infection in Denmark: a nationwide population-based cohort study. The lancet HIV. 2015;2(7):e288–e98.

12. de Paula HHS, Ferreira ACG, Caetano DG, Delatorre E, Teixeira SLM, Coelho LE, et al. Reduction of inflammation and T cell activation after 6 months of cART initiation during acute, but not in early chronic HIV-1 infection. Retrovirology. 2018;15(1):1–11.

13. McLaughlin MM, Ma Y, Scherzer R, Rahalkar S, Martin JN, Mills C, et al. Association of Viral Persistence and Atherosclerosis in Adults With Treated HIV Infection. JAMA Netw Open. 2020;3(10):e2018099.

14. Turcotte I, El-Far M, Sadouni M, Chartrand-Lefebvre C, Filali-Mouhim A, Fromentin R, et al. Association Between the Development of Subclinical Cardiovascular Disease and Human Immunodeficiency Virus (HIV) Reservoir Markers in People With HIV on Suppressive Antiretroviral Therapy. Clin Infect Dis. 2023;76(7):1318–21.

15. Morgan E, Taylor HE, Ryan DT, D’Aquila R, Mustanski B. Systemic inflammation is elevated among both HIV-uninfected and-infected young men who have sex with men. AIDS (London, England). 2019;33(4):757.

16. Lama JR, Brezak A, Dobbins JG, Sanchez H, Cabello R, Rios J, et al. Design strategy of the Sabes Study: diagnosis and treatment of early HIV infection among men who have sex with men and transgender women in Lima, Peru, 2013–2017. American journal of epidemiology. 2018;187(8):1577–85.

17. Lama JR, Ignacio RAB, Alfaro R, Rios J, Cartagena JG, Valdez R, et al. Clinical and immunologic outcomes after immediate or deferred antiretroviral therapy initiation during primary human immunodeficiency virus infection: the Sabes randomized clinical study. Clinical Infectious Diseases. 2021;72(6):1042–50.

18. Grebe E, Facente SN, Bingham J, Pilcher CD, Powrie A, Gerber J, et al. Interpreting HIV diagnostic histories into infection time estimates: analytical framework and online tool. BMC infectious diseases. 2019;19(1):1–10.

19. Nabatanzi R, Bayigga L, Cose S, Rowland Jones S, Joloba M, Canderan G, et al. Monocyte dysfunction, activation, and inflammation after long-term antiretroviral therapy in an African Cohort. The Journal of infectious diseases. 2019;220(9):1414–9.

20. Brites-Alves C, Luz E, Netto EM, Ferreira T, Diaz RS, Pedroso C, et al. Immune activation, proinflammatory cytokines, and conventional risks for cardiovascular disease in HIV patients: a case-control study in Bahia, Brazil. Frontiers in immunology. 2018;9:1469.

21. Hoenigl M, Moser CB, Funderburg N, Bosch R, Kantor A, Zhang Y, et al. Soluble urokinase plasminogen activator receptor is predictive of non-aids events during antiretroviral therapy–mediated viral suppression. Clinical Infectious Diseases. 2019;69(4):676–86.

22. Siedner MJ, Bwana MB, Asiimwe S, Amanyire G, Musinguzi N, Castillo-Mancilla J, et al. Timing of antiretroviral therapy and systemic inflammation in sub-Saharan Africa: results from the META longitudinal cohort study. The Journal of infectious diseases. 2019;220(7):1172–7.

23. Babu H, Ambikan AT, Gabriel EE, Svensson Akusjärvi S, Palaniappan AN, Sundaraj V, et al. Systemic inflammation and the increased risk of inflamm-aging and age-associated diseases in people living with HIV on long term suppressive antiretroviral therapy. Frontiers in immunology. 2019;10:1965.

24. Subramanya V, McKay HS, Brusca RM, Palella FJ, Kingsley LA, Witt MD, et al. Inflammatory biomarkers and subclinical carotid atherosclerosis in HIV-infected and HIV-uninfected men in the Multicenter AIDS Cohort Study. PloS one. 2019;14(4):e0214735.

25. Tarantino G, Costantini S, Citro V, Conforti P, Capone F, Sorice A, et al. Interferon-alpha 2 but not Interferon-gamma serum levels are associated with intramuscular fat in obese patients with nonalcoholic fatty liver disease. Journal of translational medicine. 2019;17(1):1–14.

26. Paul RF, Hassan M, Nazar HS, Gillani S, Afzal N, Qayyum I. Effect of body mass index on serum leptin levels. J Ayub Med Coll Abbottabad. 2011;23(3):40–3.

27. Deshmane SL KS, Amini S, Sawaya BE. Monocyte Chemoattractant Protein-1 (MCP-1): An Overview. J Interferon Cytokine Res. 2009;29(6):313–26.

28. Luster AD RJ. Biochemical characterization of a gamma interferon-inducible cytokine (IP-10). J Exp Med. 1987;166(4):1084–7.

29. Shi C, Pamer, E. Monocyte recruitment during infection and inflammation. Nat Rev Immunol. 2011;11:762–74.

30. Bordoni V, Sacchi A, Casetti R, Cimini E, Tartaglia E, Pinnetti C, et al. Impact of ART on dynamics of growth factors and cytokines in primary HIV infection. Cytokine. 2020;125:154839.

31. Pérez-Pérez A S-JF, Vilariño-García T, Sánchez-Margalet V. Role of Leptin in Inflammation and Vice Versa. Int J Mol Sci. 2020;21(16):5887. Published 2020 Aug 16. doi:10.3390/ijms21165887. Role of Leptin in Inflammation and Vice Versa. Int J Mol Sci. 2020;21(16).

32. Tiliscan C, Aramă V, Mihăilescu R, Munteanu DI, Streinu-Cercel A, Ion DA, et al. Leptin expression in HIV-infected patients during antiretroviral therapy. Germs. 2015;5(3):92.

33. Achur RN, Freeman WM, Vrana KE. Circulating cytokines as biomarkers of alcohol abuse and alcoholism. Journal of Neuroimmune Pharmacology. 2010;5(1):83–91.

34. Leclercq S, Cani PD, Neyrinck AM, Stärkel P, Jamar F, Mikolajczak M, et al. Role of intestinal permeability and inflammation in the biological and behavioral control of alcohol-dependent subjects. Brain, behavior, and immunity. 2012;26(6):911–8.

35. Tagarro A, Chan M, Zangari P, Ferns B, Foster C, De Rossi A, et al. Early and highly suppressive ART are main factors associated with low viral reservoir in european perinatally HIV infected children. Journal of acquired immune deficiency syndromes (1999). 2018;79(2):269.

36. McComsey GA, Kitch D, Daar ES, Tierney C, Jahed NC, Melbourne K, et al. Inflammation markers after randomization to abacavir/lamivudine or tenofovir/emtricitabine with efavirenz or atazanavir/ritonavir: ACTG A5224 s, A5202 substudy. AIDS (London, England). 2012;26(11):1371.

37. Cleary PD, Van Devanter N, Rogers TF, Singer E, Shipton-Levy R, Steilen M, et al. Behavior changes after notification of HIV infection. American journal of public health. 1991;81(12):1586–90.

38. Elias-Smale SE, Kardys I, Oudkerk M, Hofman A, Witteman JC. C-reactive protein is related to extent and progression of coronary and extra-coronary atherosclerosis; results from the Rotterdam study. Atherosclerosis. 2007;195(2):e195–e202.

39. Laaksonen DE, Niskanen L, Nyyssönen K, Punnonen K, Tuomainen T-P, Salonen JT. C-reactive protein in the prediction of cardiovascular and overall mortality in middle-aged men: a population-based cohort study. European heart journal. 2005;26(17):1783–9.

